# Stimulant Craving and Drug Use Dynamics: A Cross-Lagged Residual Dynamic Structural Equation Modeling Study

**DOI:** 10.64898/2026.05.09.26352809

**Authors:** Ramin Mojtabai, Ryoko Susukida, Trang Quynh Nguyen, Mehdi Farokhnia, Lorenzo Leggio, Cecilia Bergeria, Smita Prasad, Kelly E. Dunn, Masoumeh Amin-Esmaeili

## Abstract

**Aims:** To examine the longitudinal dynamic interactions of craving and drug use in the course of treatment of stimulant use disorders.

**Design:** Cross-lagged residual dynamic structural equation modeling (R-DSEM) was used to examine the reciprocal (bidirectional) longitudinal associations between craving and drug use.

**Setting:** Pooled data from 11 randomized controlled trials of pharmacotherapies for methamphetamine and cocaine use disorders in the United States sponsored by the National Institute on Drug Abuse.

**Participants:** 1,936 adults with cocaine or methamphetamine use disorder.

**Measurements:** Craving was measured using Brief Substance Craving Scale (BSCS), drug use was measured using Timeline Followback and urine drug screen (UDS).

**Findings:** Craving and stimulant drug use were dynamically associated over time (within-person association). Daily craving significantly predicted drug use in subsequent days (estimate=0.092, 95% credible interval [CrI]=0.081, 0.103 for self-reported drug use and estimate=0.081, 95% CrI=0.069, 0.095 for UDS-ascertained drug use). In turn, drug use predicted subsequent craving (estimate=0.361, 95% CrI=0.325, 0.398 and estimate=0.060, 95% CrI=0.028, 0.094, respectively). There was substantial between-person heterogeneity in these cross-lagged effects, as reflected in the coefficients of variation ranging from 0.78 to 2.88.

**Conclusions:** There is a bidirectional interaction between stimulant drug craving and drug use. The heterogeneity in the interaction of craving with stimulant drug use may partly explain between-person variability in responses to anti-craving medications in treatment of stimulant use disorders.

## INTRODUCTION

Drug craving has long been recognized as a key symptom in addictive behaviors, including stimulant use disorders, going back to early descriptions of these conditions [1, 2]. More recent research suggests that craving is intrinsically associated with other outcomes of stimulant use disorders, including return to drug use and remission [3–12]. Moreover, pharmacotherapeutic candidates for stimulant use disorders have often been selected based on their potential anti-craving properties [13, 14].

The central role of craving in stimulant use disorders have motivated researchers to examine the dynamic associations between drug use and craving for these drugs in the course of treatment for stimulant drug use disorders [6, 15–17]. However, with rare exceptions [6, 18], these studies did not examine possible reciprocal (bidirectional) associations between drug craving and drug use. An Ecological Momentary Assessment (EMA) study by Preston and colleagues [6] found higher levels of cocaine craving during periods of cocaine use in the course of treatment. The study also found that craving increased every hour in the 5-hour span before an episode of drug use. Another study by Serre and colleagues also used EMA to examine the reciprocal associations between craving and drug use [18]. However, the study only found a unidirectional association between craving intensity and the primary substance use at the next assessment. However, the study sample comprised a mix of substance use disorders and included only 7 participants with a stimulant (cocaine) use disorder.

A reciprocal associations between stimulant drug use and craving is plausible based on evidence from both clinical and laboratory studies suggesting that craving can be induced by exposure to stimulants [19–21]. Nonetheless, the association between stimulant craving and use may be variable across individuals. The underlying mechanisms and correlates of stimulant use disorders are heterogeneous [22, 23] and there is emerging evidence of considerable inter-individual variability in the association of cue exposure and exposure to the drug with cocaine craving [21, 24]. The inter-individual variability in the association of stimulant craving and stimulant use has potential implications for targeting anti-craving treatment to individuals who might benefit more from such treatments. However, this variability in the association of stimulant craving with stimulant use has not been systematically examined.

In this study, we used data from 11 past randomized controlled trials (RCTs) of cocaine use disorder (6 RCTs) or methamphetamine use disorder (5 RCTs) to examine the dynamic reciprocal relationship between stimulant drug craving and drug use in the course of RCTs. We used the novel method of residual dynamic structural equation modeling (R-DSEM) that is specifically designed for intensive longitudinal data (i.e., data collected frequently in a relatively short period of time) [25, 26].

In previous studies using these pooled RCT data, we identified 3 trajectories of craving in the course of RCTs for stimulant use disorders [27, 28]. These trajectories were strongly correlated with trajectories of stimulant use during treatment and with other outcomes of stimulant use disorders. The trajectory groups that experienced the greatest reduction in craving also experienced the greatest reduction in drug use and the greatest improvement in other health-related and social outcomes. These analyses, however, did not examine the dynamic time-varying associations between craving and drug use or the inter-individual heterogeneity in this relationship. In the present study, we used intensive longitudinal data collected daily or several times a week from RCT participants to examine the cross-lagged relationship between craving and both self-reported and urine drug screen (UDS)-ascertained stimulant use.

## METHOD

### Sample

Data were drawn from a harmonized dataset of 11 National Institute on Drug Abuse (NIDA)-sponsored RCTs of pharmacotherapies for stimulant use disorders (N=1,936), including 6 RCTs for cocaine use disorder (n=1,070) and 5 RCTs for methamphetamine use disorder (n=866). Among other eligibility criteria, participants had to have a DSM-IV diagnosis of cocaine or methamphetamine dependence to qualify for the cocaine or methamphetamine RCTs, respectively [29]. The medication tested in each RCT and other characteristics of the studies are summarized in Appendix A and described in our previous reports [27, 28]. All RCTs had a placebo arm, were double-blind and between-subjects, and all participants received cognitive behavioral therapy as an adjunct.

These studies were selected from a pool of 44 pharmacotherapy RCTs for cocaine or methamphetamine use disorder available, as fully anonymized data, on the NIDA Data Share site (https://datashare.nida.nih.gov/), accessed on April 5, 2023. Studies were selected if they were (1) efficacy RCTs comparing an active medication with a placebo for treatment of cocaine or methamphetamine use disorder, and (2) included the Brief Substance Craving Scale (BSCS) for assessment of drug craving (see below). Pilot, safety, and drug interaction studies were not included. The selected RCTs employed similar designs and collected almost identical measurements, justifying pooling of their samples.

Application of R-DSEM relies on a sufficient number of assessment points to provide reliable estimates of the autoregressive terms and for the models to converge [26, 30]. In their simulation studies, Gistelinck and colleagues observed that R-DSEM does not perform well when the number of time points is smaller than 10 [31]. Therefore, we limited our sample to participants with at least 10 craving assessments in the course of RCT. Additionally, participants with no variability in craving based on within person variances were excluded as these cases do not contribute to estimation and may negatively impact R-DSEM model estimates. After these exclusions, 1,212 participants remained in the analytic sample for self-reported drug use.

The sample for the analysis of UDS-ascertained drug use and craving was additionally limited to participants with at least 10 UDS assessments. As noted, R-DSEM has been shown not to perform well when the number of time points is smaller than 10 [31]. As a result, data from 4 trials with fewer than 10 UDS assessments (NIDA-CTO-0001, NIDA-CTO-0007, NIDA-CSP-1025, NIDA-CSP-1026) and individual-level data from participants in other RCTs with less than 10 UDS assessments were excluded, resulting in a sample size of 622 for the analysis of UDS-ascertained drug use. We examined the potential impact of these exclusions on generalizability of the results in a set of sensitivity analyses (see Statistical analysis).

All RCTs were approved by the respective institutions’ ethical boards, and this secondary analysis was deemed exempt from review by the Tulane University Institutional Review Board.

### Measures

Methods used for harmonization of the RCT data have been previously described [32]. Briefly, all core measures, including demographic data, psychosocial measures, craving assessments, and measures of drug use, were harmonized across RCTs for pooled analyses.

Craving for stimulant drugs was assessed using the multi-item BSCS, administered weekly in all RCTs. The BSCS is based on the drug craving section of the *State of Feelings and Cravings Questionnaire* developed by Mezinskis and colleagues [33, 34]. The questions ask about the intensity, frequency, and duration of craving in the past 24 hours for the primary and secondary drugs. For this study, we only used BSCS ratings for the primary drug (cocaine in cocaine RCTs and methamphetamine in methamphetamine RCTs). Intensity was measured on a Likert scale ranging from “not at all” (=0) to “extreme” (=4); frequency was measured on a scale from “never” (=0) to “almost constantly” (=4); and duration was measured on a scale from “none at all” (=0) to “very long” (=4). Consistent with the standard scoring procedure for computing a BSCS summary score, the scores on the 3 items were summed up (range=0-12) [35]. Past research has established the internal consistency and construct validity of the BSCS, as well as its sensitivity to treatment effect [33, 34], and its utility in clinical research [36, 37]. Mezinskis and colleagues reported a high correlation between BSCS rating of cocaine craving and self-reported use of cocaine (Pearson *r*=.50) [33].

Primary drug use was ascertained by self-report and UDS 3 times a week in all 11 RCTs per protocol. Self-reported drug use was assessed using the Timeline Followback (TLFB) [38] to retrospectively assess use of the primary drug since the last assessment. Self-reports and UDS ascertained use corresponded strongly. Fully 93% of those who reported use of a stimulant drug on a certain day and were also tested for the drug on that day had a positive UDS. Among those who did not report stimulant drug use on the day when they had a positive UDS, 73% reported a drug in the previous 4 days, which is plausible given that UDS reflects drug use over the past several days (typically 2–4 days) [39].

Sociodemographic characteristics, including sex, age, and race/ethnicity were assessed using a standardized questionnaire.

### Statistical analysis

We used R-DSEM implemented in *Mplus* version 9 to examine cross-lagged associations between craving and drug use [25, 26]. R-DSEM integrates multilevel modeling, time-series analysis, and structural equation modeling within a unified Bayesian framework, allowing the simultaneous estimation of within-person temporal dynamics and between-person heterogeneity in those dynamics (Figure 1). R-DSEM is preferable to standard DSEM when stationarity assumptions are violated, such as in these data, where previous research has documented temporal trends in both drug use and craving in the course of the RCTs [27, 28].

**Figure 1:**
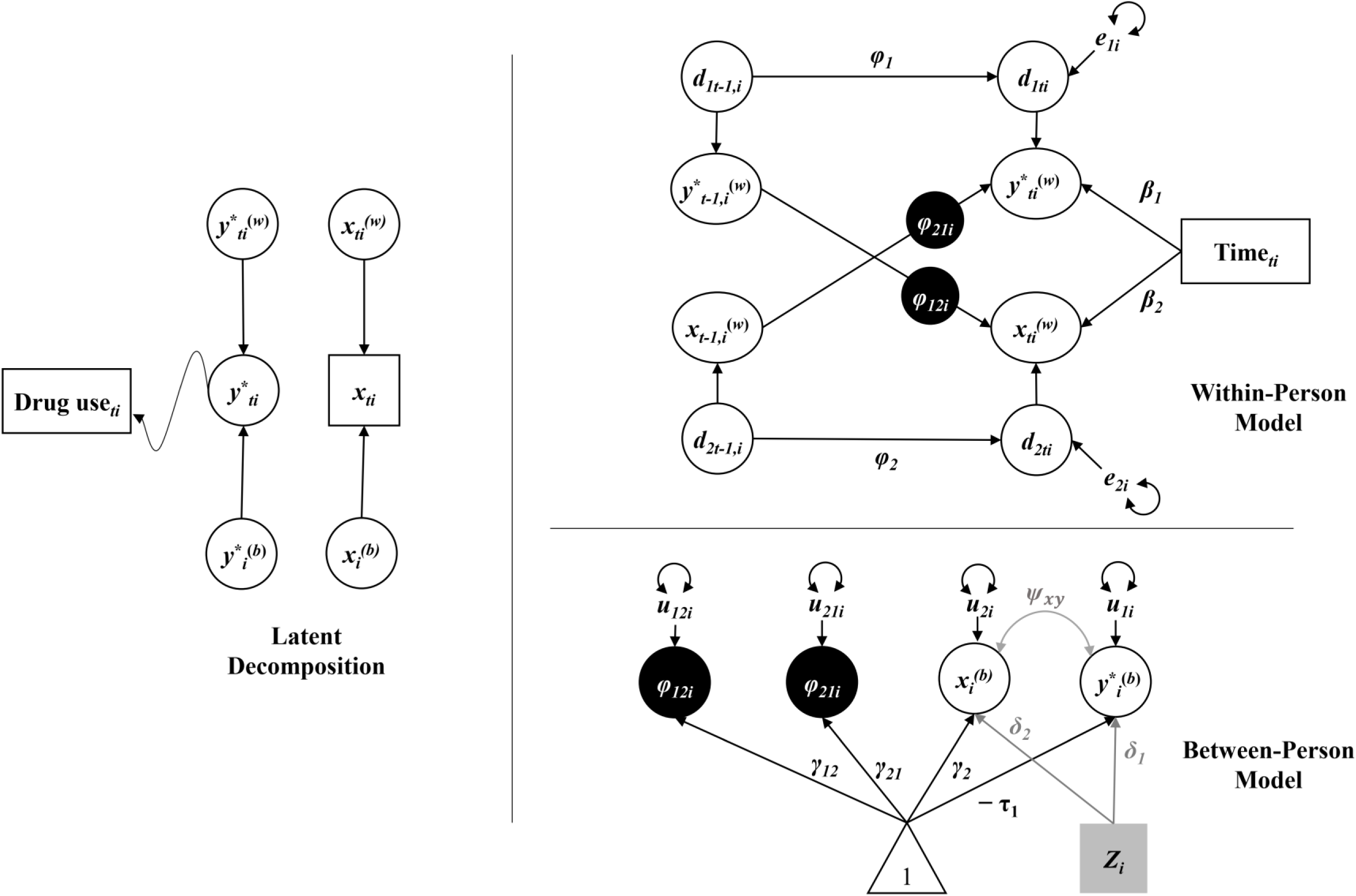
Residual Dynamic Structural Equation Model (R-DSEM) for stimulant drug use and craving. The diagram illustrates the latent decomposition, within-person, and between-person components of the R-DSEM. **Note.** Observed variables are shown as squares and latent variables as circles. Single-headed arrows represent regression paths, and curved double-headed arrows represent variances. The binary observed stimulant drug use indicator is linked to a latent response variable *y^*^_ti_* through a probit threshold τ_1_, such that Pr(Drug use_ti_ = 1) = Φ(y^*^_ti_. Both *y^*^_ti_* and craving *x*_*ti*_ are decomposed into within-person (*W*) and between-person (*b*) components. At the within-person level, deviations in stimulant drug use liability and craving are linked by individual-specific cross-lagged effects ф_21*i*_ (craving → subsequent drug use liability) and ф_12*i*_ (drug use liability → subsequent craving), adjusted for linear time trends β_1_ and β_2_. Residual innovation processes *d*_1*ti*_ and *d*_2*ti*_ follow autoregressive structures with parameters ф_1_ and ф_2_. At the between-person level, the latent intercept *y^*(b)^_ti_*, mean craving component *x^(b)^_i_*, and the random cross-lagged effects ф_21*i*_ and ф_12*i*_ are modeled as functions of population means and person-specific deviations. Time-invariant covariates *Z_i_*, when included, predict *y^Ȋ(b)^_ti_* and *x^(b)^_ti_* via regression coefficients δ_1_ and δ_2_ (See Appendix B for mathematical representation of the model).

R-DSEM decomposes each time-varying variable into a stable between-person component (random intercept) and a time-specific residual, and models autoregressive terms exclusively among these residuals. Further, the effect of time on the outcome variables (craving and drug use) was modeled at the within-person level. By moving the autoregressive effects to the residuals, R-DSEM improves the interpretability of temporal effects as within-person processes rather than stable between-person differences.

Cross-lagged effects of craving on subsequent drug use and vice versa are also modeled at the within-person level, capturing how deviations from an individual’s own mean on an outcome at timepoint *t* predict deviations of the other outcome at a subsequent timepoint (*t* + 1). Variables were person-mean centered so that within-person parameters reflect purely intra-individual processes. Lagged effects were specified using a first-order discrete-time approximation. Because UDS and craving assessments occurred at unequal intervals across outcomes and participants, lag lengths varied. *Mplus* corrects for unequally spaced observations via the time-interval (TINTERVAL) correction of the R-DSEM using a Kalman filter–based adjustment [26].

At the between-person level, the random effects represent differences that are modeled as latent variables. Between-person means of the random effects (*γ₁₂* and *γ₂₁* in Table 1) quantify average dynamic relationships, while their variances (Var(*u₁₂i*), Var(*u₂₁i*)) capture heterogeneity in within-person mechanisms across individuals.

**Table 1:**
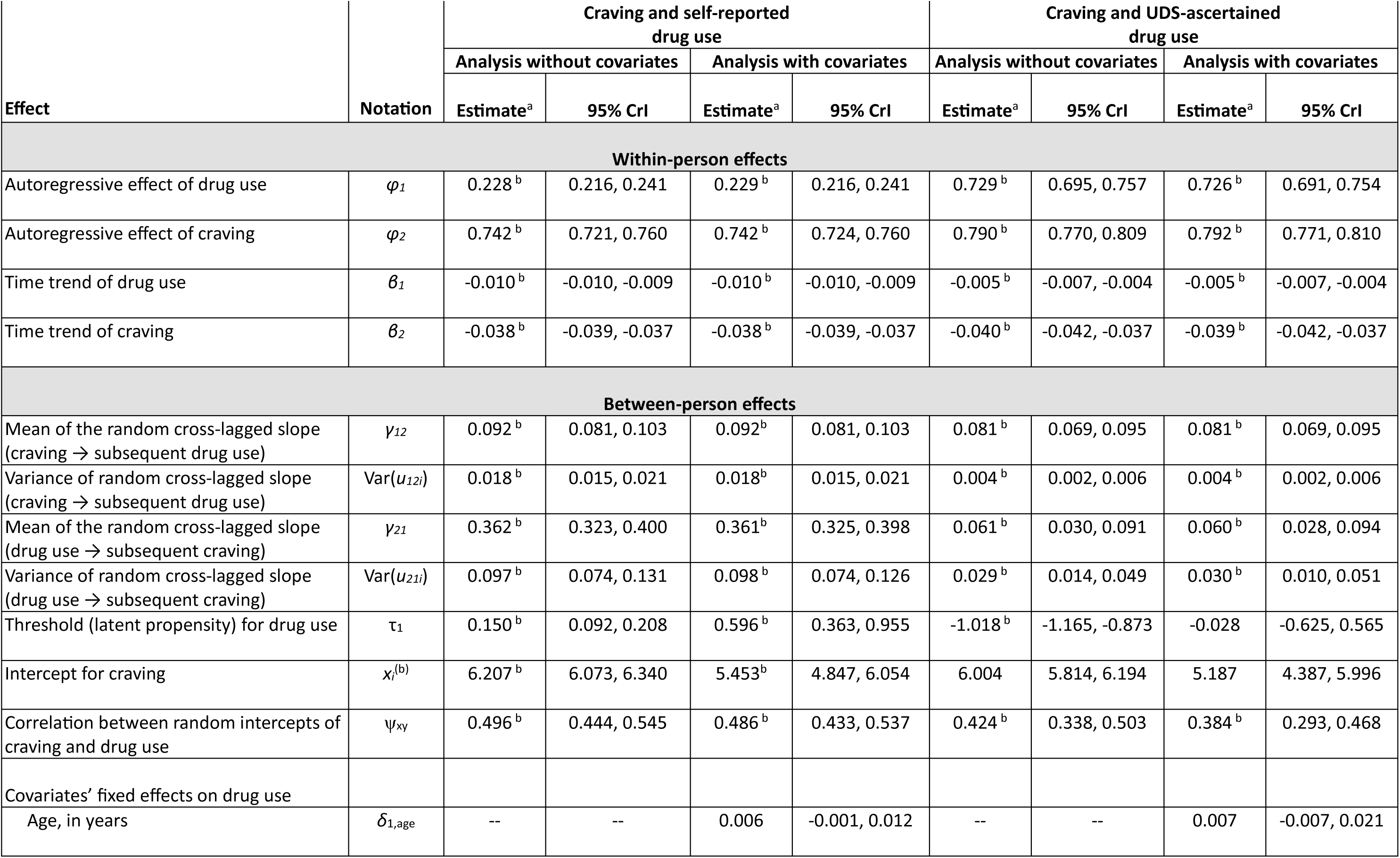
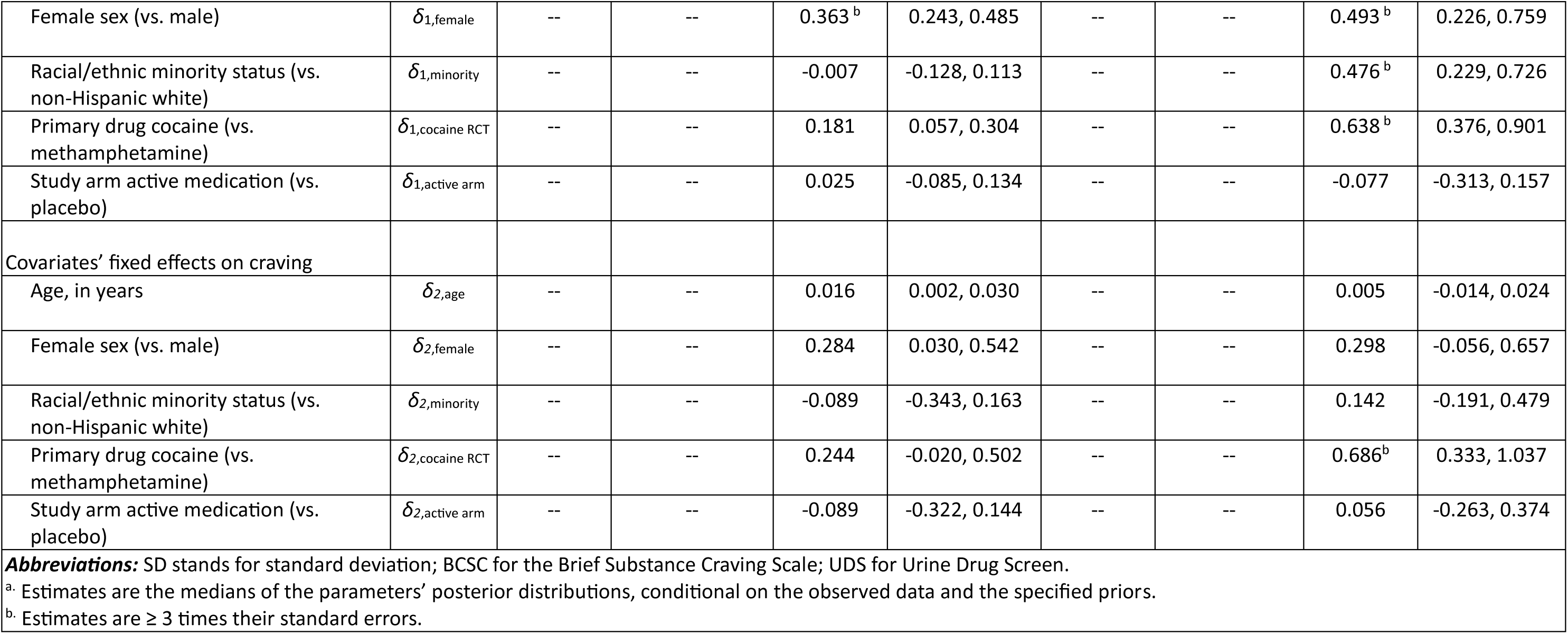
Results of the Residual Dynamic Structural Equation Modeling (R-DSEM) analyses of the relationship between craving and self-reported / urine drug screen (UDS)-ascertained drug use in the course of randomized controlled trials of pharmacotherapies for stimulant use disorders.

As our study focused on the cross-lagged effects and their potential heterogeneity, we modeled these terms as random effects. The two outcomes included in the cross-lagged models were craving (BSCS score) and the binary variables of self-reported and UDS-ascertained drug use, modeled separately. Drug use variables were modeled as categorical outcomes via a latent response formulation using a probit link, while craving was treated as continuous. Accordingly, the estimated associations with the binary drug use outcomes reflect changes in the underlying latent propensity (liability) for drug use, expressed in standard deviation (SD) units of the latent response variable rather than direct changes in observed probabilities.

We also estimated the correlation between individuals’ random intercepts for drug use and craving (ψ_xy_), representing the association between mean craving levels and mean latent propensity for drug use.

Two sets of R-DSEM models were run, one for the cross-lagged associations of self-reported drug use and craving and another, for the associations of UDS-ascertained drug use and craving. In each set, separate models with and without covariates were run. The time-invariant covariates included sex, age, race/ethnicity (non-Hispanic white vs. other), primary drug (cocaine vs. methamphetamine), and treatment arm (active medication vs. placebo). The model equation is presented and described in Appendix B.

Models were estimated using a Bayesian Markov chain Monte Carlo (MCMC) estimator, which is well-suited for complex hierarchical time-series models and unbalanced data. Weakly informative priors were specified for fixed effects and variance components to stabilize estimation while allowing the data to dominate posterior estimates. A minimum of 20,000 iterations was used, with the first half of the iterations used as burn-in. The parameters were estimated based on the second half of iterations. Convergence of the models was assessed by examining Bayesian trace plots and Potential Scale Reduction (PSR) values. PSR is based on a comparison of the parameter variations within each chain and across multiple iterations to the parameter variations across chains. Trace plots that form a relatively tight horizontal band with no large discrepancies between the chains and a stable PSR<1.05 indicate good convergence. Further iterations after reaching this PSR value are to ensure that PSR does not increase in later iterations [40]. Missing data are handled naturally under the Bayesian framework via full-information estimation, assuming data are missing at random [41].

Parameter estimates are reported as posterior medians with 95% Bayesian credible intervals (CrI). Medians, instead of means, are reported as a more robust measure of central tendency, given skewness in the posterior distribution. To ensure that the observed dynamic relationships were both statistically robust and clinically meaningful, we adopted a conservative heuristic where estimates were only considered substantive if they were at least three times their standard errors [42].

Besides parameter estimates, R-DSEM also reports the variance of random slopes as a measure of between-person heterogeneity. We further explored this heterogeneity by computing the coefficients of variation (CV) as the ratios of the standard deviations of the random slopes over corresponding parameter estimates [43, 44].

In addition to computing within-person effects for the group, we computed plausible values of these effects for each person by imputing the distribution of plausible values of latent variables, specifically the cross-lagged effects [45]. The median and 95% credible intervals for each person were calculated based on 200 imputations. The results for these estimations for the two cross-lagged effects were summarized in a set of caterpillar plots. *Mplus* 9 software with a Bayesian estimator and a two-level random effect model was used for the R-DSEM analyses.

### Sensitivity analyses

Two sets of sensitivity analyses were conducted. First, to assess potential bias in R-DSEM analyses due to exclusion of participants with less than 10 assessments, we conducted a series of Bayesian mixed effect regression models in which auto-regressive effects (effect of craving and drug use assessments on subsequent assessments of the same outcomes) and cross-lagged effect (effect of craving on subsequent drug use outcomes and vice versa) were estimated for the total sample, the sample for self-reported drug use and the sample for UDS-ascertained drug use. Linear regression was used for analyses of craving and probit regression for the binary drug use outcomes. *Stata* 19 software’s *bayes: mixed* and *bayes: meprobit* routines were used for these analyses.

In a second set of sensitivity analyses we examined the impact of combining data from different trials on the R-DSEM results. To this end we entered dummy variables for the individual RCTs as time-invariant covariates into the R-DSEM models for self-reported and UDS-ascertained drug use.

## RESULTS

The characteristics and outcome ratings of participants in the two analytic samples and total pooled RCT sample are presented in Appendices C. The samples for analyses of self-reported drug use and UDS-ascertained drug use mostly had similar sociodemographic characteristics to the total sample. The average ages in these samples ranged from 39.7 to 40.5 years, being somewhat higher in the sample for examining self-reported drug use. Across the samples, the majority of participants were male (70.1% to 72.5%) and from racial/ethnic minority groups (54.5% to 55.2%).

The average level of craving and the frequency of both self-reported and UDS-ascertained drug use were somewhat lower in the analytic samples for self-reported and UDS-ascertained drug use compared with the total sample (Appendix C).

### R-DSEM analyses

The R-DSEM models converged normally with stable PSR levels below 1.05 and satisfactory Bayesian trace plots (Appendix D). The results of the R-DSEM analyses are presented in Table 1 for models with and without covariates. Below, we describe the results of the models with covariates, as estimates were similar in models with and without covariates.

Both self-reported and UDS-ascertained drug use exhibited strong temporal stability at the within-person level, reflected in significant autoregressive effects that were stronger for UDS-ascertained drug use than for self-reported drug use (estimate =0.229, 95% credible interval [CrI]=0.216, 0.241 for self-reported drug use; estimate=0.726, 95% CrI=0.691, 0.754 for for UDS-ascertained drug use). Craving also showed significant persistence over time (estimate =0.742, 95% CrI=0.724, 0.760 in the model for self-reported drug use; estimate=0.792, 95% CrI=0.771, 0.810 in the model for UDS-ascertained drug use).

Time trends were significant and negative for self-reported and UDS-ascertained drug use as well as for craving, indicating that both drug use and craving decreased in the course of RCTs (Table 1).

The analyses also revealed significant bidirectional cross-lagged associations between craving and drug use at within-person level. Increased craving compared to the person’s mean craving level predicted an increased likelihood of subsequent self-reported drug use (estimate = 0.092, 95% CrI=0.081, 0.103) as well as subsequent UDS-ascertained drug use (estimate = 0.081, 95% CrI=0.069, 0.095).

A parameter estimate of 0.092 for craving indicates that a 1-unit increase in craving (above a person’s own average) increases the latent liability of subsequent self-reported drug use by approximately 0.09 SD units, corresponding to roughly a 3–4 percentage-point increase in the probability of drug use when the baseline probability is around 30%. Similarly, a parameter estimate of 0.081 for craving in the probit model predicting UDS-ascertained drug use indicates that a 1-unit increase in craving increases the latent liability of subsequent drug use by approximately 0.08 SD units, also corresponding to approximately a 2–3 percentage-point increase in probability of subsequent drug use when the baseline probability is around 65%.

Conversely, higher drug use propensity predicted higher subsequent levels of craving. In the model for self-reported drug use, the within-person cross-lagged effect of drug use on subsequent craving was substantial (estimate = 0.361, 95% CrI = 0.325, 0.398). In the model for UDS-ascertained drug use, the effect was smaller (estimate = 0.060, 95% CrI = 0.028, 0.094). However, both effects were significant as the 95% credible intervals did not include 0. Because the model uses a probit link, these estimates represent the change in subsequent craving per unit increase in the latent propensity of drug use. Notably, the drug use effect on craving was substantially stronger in the self-reported model than in the UDS-ascertained model.

At between-person level, participants with a higher propensity of drug use tended to have significantly higher levels of overall craving as evidenced by the correlations of mean level of drug use propensity and craving (ψ_xy_=0.486 for self-reported drug use with craving and ψ_xy_=0.384 for UDS-ascertained drug use with craving).

Moreover, the random slope variances (Table 1) indicated substantial between-person heterogeneity in both cross-lagged pathways. We quantified this heterogeneity as a median-based CV (the ratio of the standard deviation of the random slope to its posterior median) [43, 44]. CV was estimated as 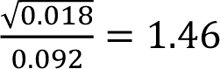 for the craving effect on subsequent self-reported drug use and 0.87 for the effect of self-reported drug use on subsequent craving. Both values indicate substantial posterior dispersion and heterogeneity in the estimates. CV’s were similarly large for the effect of craving on subsequent UDS-ascertained drug use (0.78), and the effect of UDS-ascertained drug use on subsequent craving (2.88). This heterogeneity in the association of craving and drug use outcomes was also apparent in the caterpillar plots of within-person standardized effects for individual participants (Figure 2A-D). The pink-colored credible intervals in these plots indicate individuals for whom the intervals were significantly different from 0.

**Figure 2A-D:**
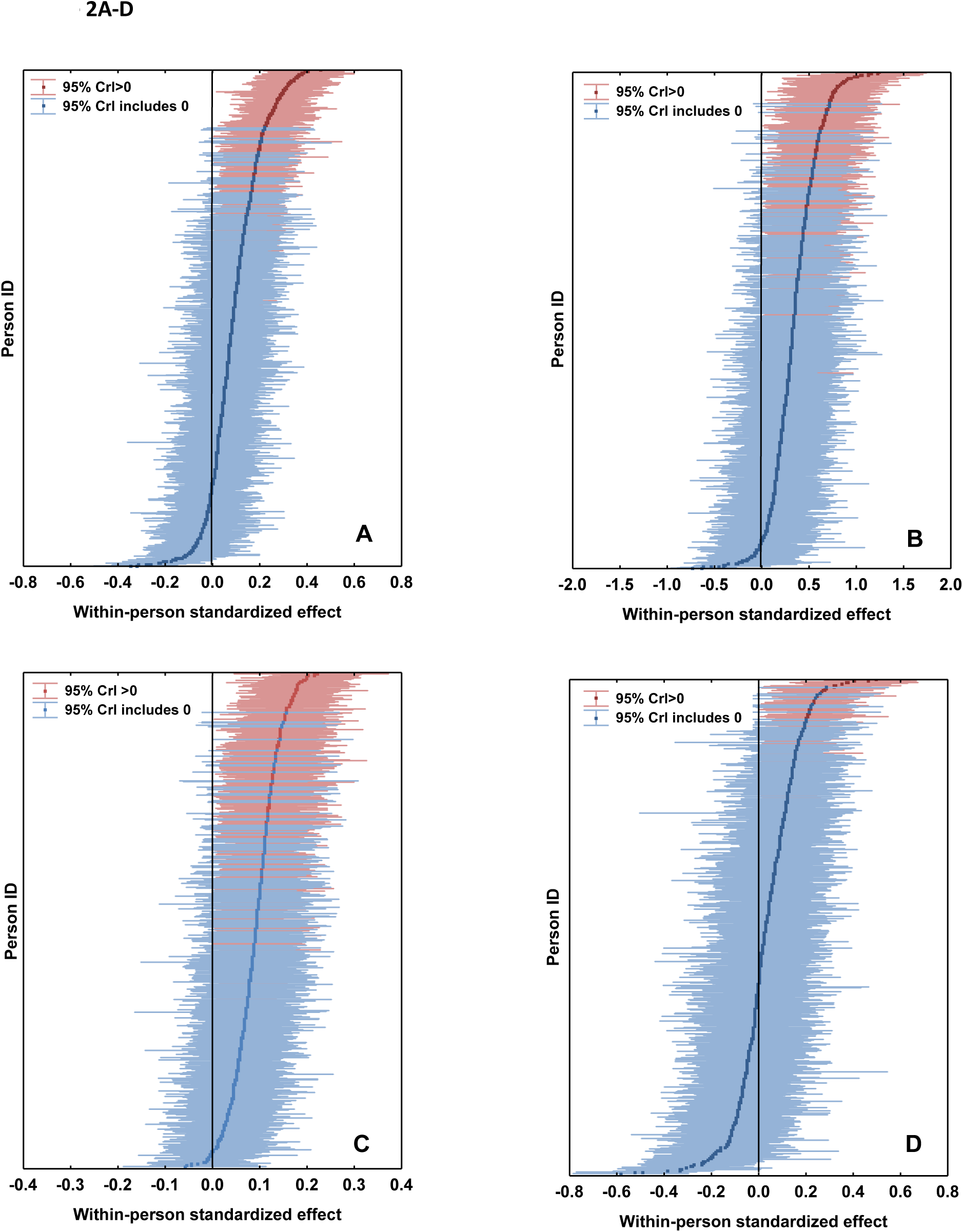
Person-specific parameter summaries of the random cross-lagged effects estimated from the Residual Dynamic Structural Equation Model (R-DSEM). Panels display posterior medians and 95% credible intervals of individual-specific cross-lagged parameters (ф_21*i*_ and ф_12*i*_). The effects shown are: (A) craving → subsequent self-reported stimulant use, (B) self-reported stimulant use → subsequent craving, (C) craving → subsequent urine drug screen (UDS)-ascertained stimulant use, and (D) UDS-ascertained stimulant use → subsequent craving.

The within-person and between-person effects in models with covariates were quite similar to those in models without covariates. Further, several of the covariates were significantly associated with the outcomes. Female sex was associated with a higher estimated propensity of both self-reported and UDS-ascertained drug use (estimate=0.363, 95% CrI=0.243, 0.485 and estimate=0.493, 95% CrI=0.226, 0.759, respectively) (Table 1). The probability of drug use was 0.38 in female participants vs. 0.30 in males for self-reported use and 0.70 vs. 0.63, respectively, for UDS-ascertained use.

Participants in cocaine RCTs also tended to have a higher estimated propensity of UDS-ascertained drug use compared to participants in methamphetamine RCTs (estimate=0.638, 95% CrI=0.376, 0.901) (Table 1). The probability of UDS-ascertained drug use was 0.34 in cocaine RCTs vs. 0.30 in methamphetamine RCTs. In addition, participants from racial/ethnic minority groups had higher UDS-ascertained drug use compared to non-Hispanic whites (estimate=0.476, 95% CrI=0.229, 0.726).

Participants in cocaine RCTs also tended to have higher levels of craving than participants in methamphetamine RCTs in analyses for UDS-ascertained drug use (estimate=0.686, 95% CrI=0.333, 1.037). The mean level of craving was 4.6 (SD=1.9) in cocaine RCTs vs. 3.7 (SD=3.7) in methamphetamine RCTs.

### Sensitivity analyses

In sensitivity analyses examining auto-regressive and cross-lagged effects using Bayesian mixed regression models, estimates of these effects were similar in the total sample and the analytic samples. Assessments of craving and drug use at each time point were significantly associated with subsequent assessments of the same outcomes. Furthermore, the cross-lagged effects indicated significant associations between assessments of craving at each time point with subsequent drug use and vice-versa (Appendix E).

In the sensitivity analysis to assess the effect of pooling RCTs we entered fixed effects for individual RCTs into the R-DSEM models. There was some variability in the average propensity of self-reported and UDS-ascertained drug use as well as craving across different RCTs (Appendix F). However, the within-person and between-person effects, including the cross-lagged estimates in models including these fixed effects were similar to the main analyses.

## DISCUSSION

This study found dynamic and reciprocal associations between craving and drug use in the course of pharmacotherapy trials for stimulant use disorders. The cross-lagged estimates suggest that elevations in craving on a given day were associated with higher probability of subsequent drug use, and drug use in turn was associated with heightened craving on subsequent days. Furthermore, the significant correlations between the mean latent propensity for drug use and craving indicate that individuals who, on average, were more likely to use cocaine or methamphetamine during the trials also tended to experience higher overall levels of craving.

These results are consistent with past research on the association between craving and subsequent drug use in the course of treatment for stimulant use [6, 15–17]. What this study adds to the extant body of research is to demonstrate that the association between craving and drug use is dynamic and bidirectional. Increased craving leads to increased likelihood of drug use, which in turn leads to higher levels of craving. This vicious cycle potentially contributes to persistence of drug use.

The use of R-DSEM also allowed us to explicitly examine the heterogeneity in the association of drug use and craving. The substantial heterogeneity in the association of drug use and craving is in line with our earlier work using group-based trajectory modeling [27, 28], which identified three distinct trajectory groups in the course of RCTs: a group with consistently high craving and drug use, a second group with sharply declining craving and both self-reported and UDS-ascertained drug use, and a third group with declining craving but a less drastic decline in drug use. Both the results of the present study and the previous findings from trajectory analysis suggest that while drug use and craving are associated in the course of treatment, this association is not uniform across different individuals and in some individuals, this association is weak or non-existent. This variability may partly explain variability in response to candidate medication treatments for stimulant use disorders, most of which have anti-craving properties [46, 47].

The dynamic patterns of drug craving in the course of treatment likely have implications for treatments for other substance use disorders as well. A recent EMA study of 39 individuals with alcohol, tobacco, cannabis, or opioid use disorder identified associations between the craving course (i.e., change in intensity) and inertia (i.e., the tendency to persist) in the first 14 days of treatment with long-term (5 years) outcome of treatment [48]. The study also found that increase in craving intensity was significantly associated with a greater likelihood of primary drug use approximately four hours later. However, the association of craving and drug use was not predictive of long-term outcome. These results are not directly comparable to our results as the study did not include any participants with stimulant use disorders.

The observed differences in stability of UDS-ascertained and self-reported drug use as well as their cross-lagged associations with craving are notable. UDS reflects drug use over the past several days (typically 2–4 days) [39], whereas self-report captures daily use. Consequently, temporal stability is naturally higher for UDS-ascertained use. In contrast, the larger cross-lagged effects of self-reported drug use on subsequent craving suggest that recent drug-taking behavior may act as a cue or trigger that intensifies craving.

Several limitations of this study should be noted. First, despite the relatively large sample size, only a limited number of covariates and complex associations among variables could be examined within the R-DSEM due to the relatively small number of assessment points for UDS and craving. Second, because of the requirements of the R-DSEM model, the samples had to be restricted to participants with an adequate number of assessment points and sufficient variability within drug use measures. However, the demographic compositions of these reduced samples were for the most part similar to the composition of the total sample. Furthermore, sensitivity analyses using Bayesian mixed regression modeling which does not have restrictions with regard to minimum number of assessment points found mostly very similar effect estimates for auto-regressive and cross-lagged effects between the restricted analytic samples and the total sample, suggesting that use of restricted samples in the R-DSEM analyses is unlikely to have biased population estimates of these effects. Lastly, substance use disorders are associated with significant adverse social and health outcomes. This study only focused on drug use and drug craving—two outcomes that were repeatedly measured in the course of the RCTs. Future studies with repeated measures of other outcomes can further explore dynamic relationships between these outcomes and other health and social outcomes. Furthermore, although this study relied on a validated and widely used self-report measure of craving, ongoing research is examining craving-related biomarkers, such as the fMRI-based neurobiological craving signature [49]. Future studies could investigate the dynamic relationship between drug use and fMRI-based neurobiological craving signatures and/or other biomarkers.

In the context of these limitations, this study is among the first to apply R-DSEM in the analysis of RCT data in the addiction field. The study provides further evidence supporting the dynamic interplay of craving and drug use in the course of pharmacotherapeutic RCTs in people with stimulant use disorders. The study also highlights the heterogeneity in the association of craving with drug use. Further exploration of this heterogeneity may help identify those people whose drug use is more or less strongly linked to craving in the course of treatment, and thereby identify potential subgroups with differential response to anti-craving pharmacotherapies [28, 46].

## Data Availability

The analyses are based on publicly available deidentified data on the NIDA Data Share site (https://datashare.nida.nih.gov/).

## Appendices

## Appendix A: Characteristics of the included randomized controlled trials (RCTs) of pharmacotherapies for stimulant use disorders

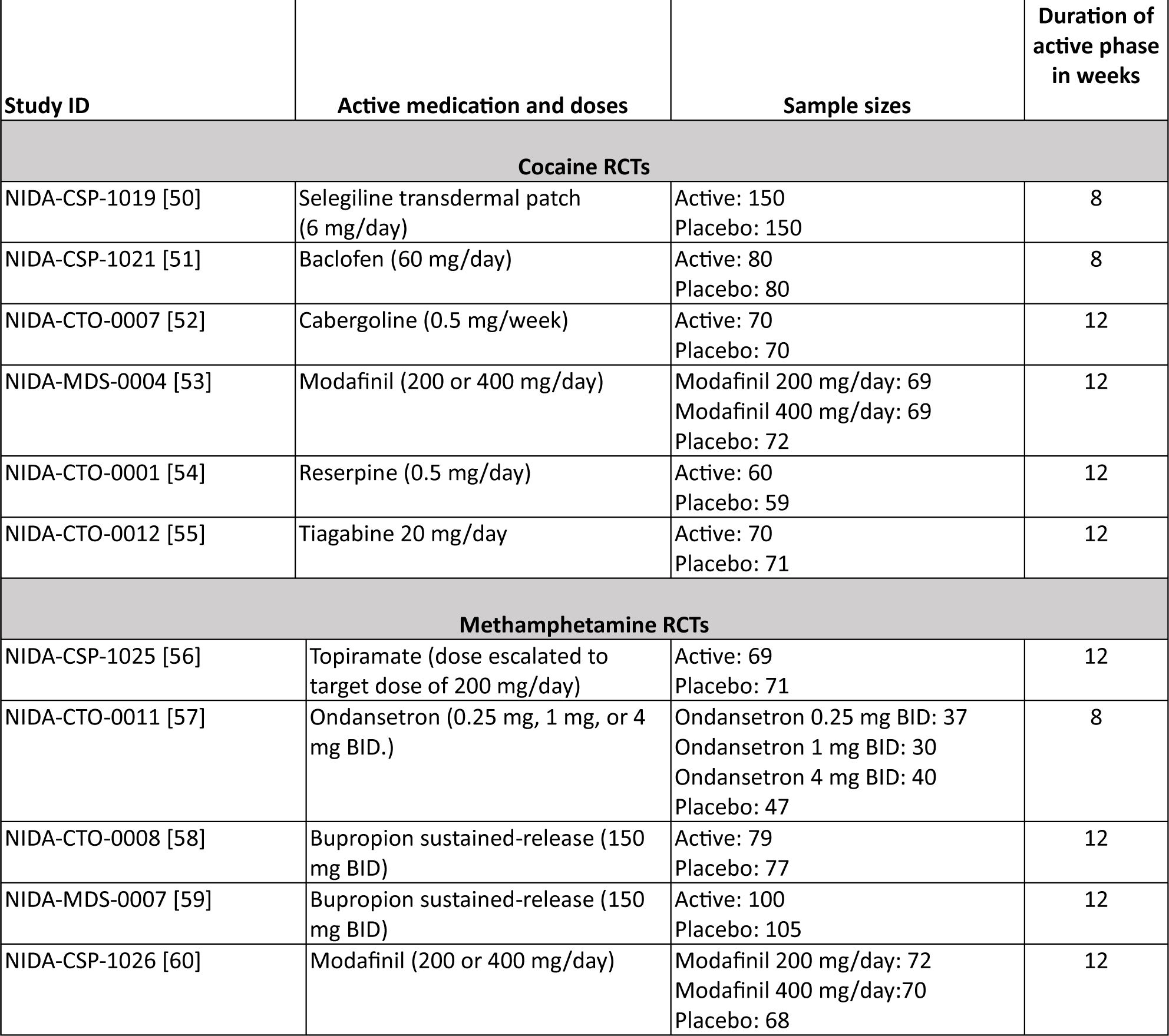

## Appendix B: Mathematical description of the Residual Dynamic Structural Equation model (R-DSEM)

The R-DSEM model for these analyses comprises three components: (1) the latent decomposition of the outcomes: drug use (binary), and craving (continuous); (2) a within-person component with random slopes for the cross-lagged effect as well as autoregressive effects on residual innovations; and (3) a between-person component for the intercept of the probit model for the binary outcomes and for the continuous outcome, and the means and variances of the within-person random cross-lagged slopes. Time-invariant covariates are added to the between-person component in models including covariates. These components are described below:

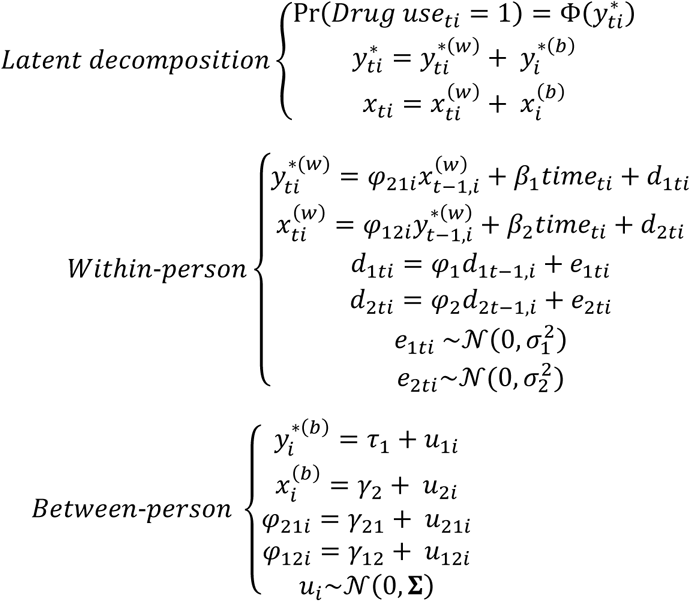

with the binary drug use indicator denoted by *y^*^_ti_*, and the propensity (liability) of drug use defined as *Pr(Drug use_ti_ = 1) = Φ(y^*^_ti_)*. The latent response is decomposed into within- and between-person components as *y^*^_ti_ = y^*(w)^_ti_ + y^*(b)^_ti_*, where *y^*(b)^_ti_ = −τ_1_ + u_1i_* represents the person-specific deviation from the population threshold τ_1_. Craving is decomposed similarly as *x_ti_ = x^(w)^_ti_ + x^(b)^_i_*. Within-person dynamics are modeled via

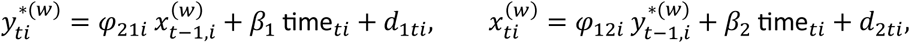

where ф_21*i*_ and ф_12*i*_ are the cross-lagged effects of craving on subsequent stimulant drug use liability and of stimulant drug use liability on subsequent craving, respectively. The time trends are indexed by β_1_and β_2_, and the residual innovation processes *d*_1*ti*_ and *d*_2*ti*_ capture the autoregressive effects for drug use (*d*_1*ti*_ = ф_1_ *d*_1,*t*−1,*i*_ + *e*_1*ti*_) and craving (*d*_2*ti*_ = ф_2_ *d*_2,*t*−1,*i*_ + *e*_2*ti*_), respectively, for a 1-day lag, adjusted for unequally spaced observations via the time-interval correction. The terms σ_1_^2^and σ_2_^2^ represent the estimated variances of the residuals.

At the between-person level, the person-specific latent intercept *y^*(b)^_ti_*, the mean craving component *x_i_^(b)^_i_*, and the individual cross-lagged effects ф_21i_ and ф_12*i*_ are modeled, along with *u*_*i*_, where *u*_*i*_ = (*u*_1*i*_, *u*_2*i*_, *u*_21*i*_, *u*_12*i*_) is a vector of person-specific random effects. In models with covariates, the vector of covariates is included as predictors of *y^*(b)^_ti_* and *x^(b)^_i_* at the between-person level, replacing the sub-models for the intercept *y^*(b)^_ti_* and *x^(b)^_i_* with *y^*(b)^_ti_* = τ_1_ + δ^T^_1_Z_i_ + u_1i_ and *x_i_^(b)^_i_* = γ_2_ + δ^T^_2_Z_i_ + u_2i_, respectively, where *Z*_*i*_ represents the vector of between-person covariates and δ_1_ and δ_2_ are the corresponding regression coefficients.

Separate models are estimated for self-reported drug use and urine drug screen (UDS)-ascertained drug use. As such, drug use in the above model is self-reported use in the model for self-reported drug use and UDS-ascertained drug use in the model for this outcome.

## Appendix C: Characteristics of the participants in the total sample and analytic samples

| **Appendix C: Characteristics of the participants in the total sample and analytic samples.** |  |  |  |
| --- | --- | --- | --- |
| **Characteristics** | **Total sample**<br>**(n=1,936)** | **Sample for the analysis of**<br>**self-reported drug use and**<br>**craving**<br>**(n=1,212)** | **Sample for the analysis of**<br>**UDS-ascertained drug use**<br>**and craving**<br>**(n=622)** |
| **Demographic characteristics of participants** |  |  |  |
|  | **Mean (SD)** | **Mean (SD)** | **Mean (SD)** |
| Age in years | 39.7 (8.6) | 40.5 (8.5)<sup>a</sup> | 40.0 (8.8) |
|  | **N (%)** | **N (%)** | **N (%)** |
| Sex |  |  |  |
| Male | 1,358 (70.1) | 852 (70.3) | 451 (72.5) |
| Female | 578 (29.9) | 360 (29.7) | 171 (27.5) |
| Race/ethnicity |  |  |  |
| Non-Hispanic White | 878 (45.5) | 543 (44.9) | 281 (45.4) |
| Non-Hispanic Black | 677 (35.1) | 435 (36.0) | 210 (33.9) |
| Hispanic | 239 (12.4) | 148 (12.2) | 93 (15.0) |
| Other | 136 (7.1) | 83 (6.9) | 35 (5.7) |
| **Participant outcomes over the course of the study** |  |  |  |
|  | **Mean (SD)** | **Mean (SD)** | **Mean (SD)** |
| Craving (BCSC) | 4.8 (2.2) | 4.4 (2.0)<sup>a</sup> | 4.2 (2.0)<sup>a</sup> |
| Frequency of self-reported drug use | 0.36 (0.27) | 0.33 (0.25)<sup>a</sup> | 0.26 (0.21)<sup>a</sup> |
| Frequency of UDS-ascertained drug use | 0.75 (0.30) | 0.75 (0.29)<sup>a</sup> | 0.65 (0.28)<sup>a</sup> |
| **Study Characteristics** |  |  |  |
|  | **N (%)** | **N (%)** | **N (%)** |
| Primary drug |  |  |  |
| Cocaine | 1,070 (55.3) | 676 (55.8) | 363 (58.4) |
| Methamphetamine | 866 (44.7) | 536 (44.2) | 259 (41.6) |
| Study arm |  |  |  |
| Active drug | 1,067 (55.1) | 694 (57.3) | 363 (58.4) |
| Placebo | 869 (44.9) | 518 (42.7) | 259 (41.6) |
| **Abbreviations:** SD stands for standard deviation. |  |  |  |
| <sup>a</sup> Differences with the total sample statistically significant at p<0.05 based on binomial test for probabilities and ordinary least square regression for continuous variables. |  |  |  |

## Appendix D: Panels display Markov chain Monte Carlo (MCMC) trace plots of the cross-lagged parameters: craving → subsequent self-reported stimulant use (panel A), self-reported stimulant use → subsequent craving (panel B), craving → subsequent urine drug screen (UDS)-ascertained stimulant drug use (panel C), and UDS-ascertained stimulant drug use → subsequent craving (panel D). Trace plots illustrate adequate mixing and convergence of the posterior distributions following burn-in

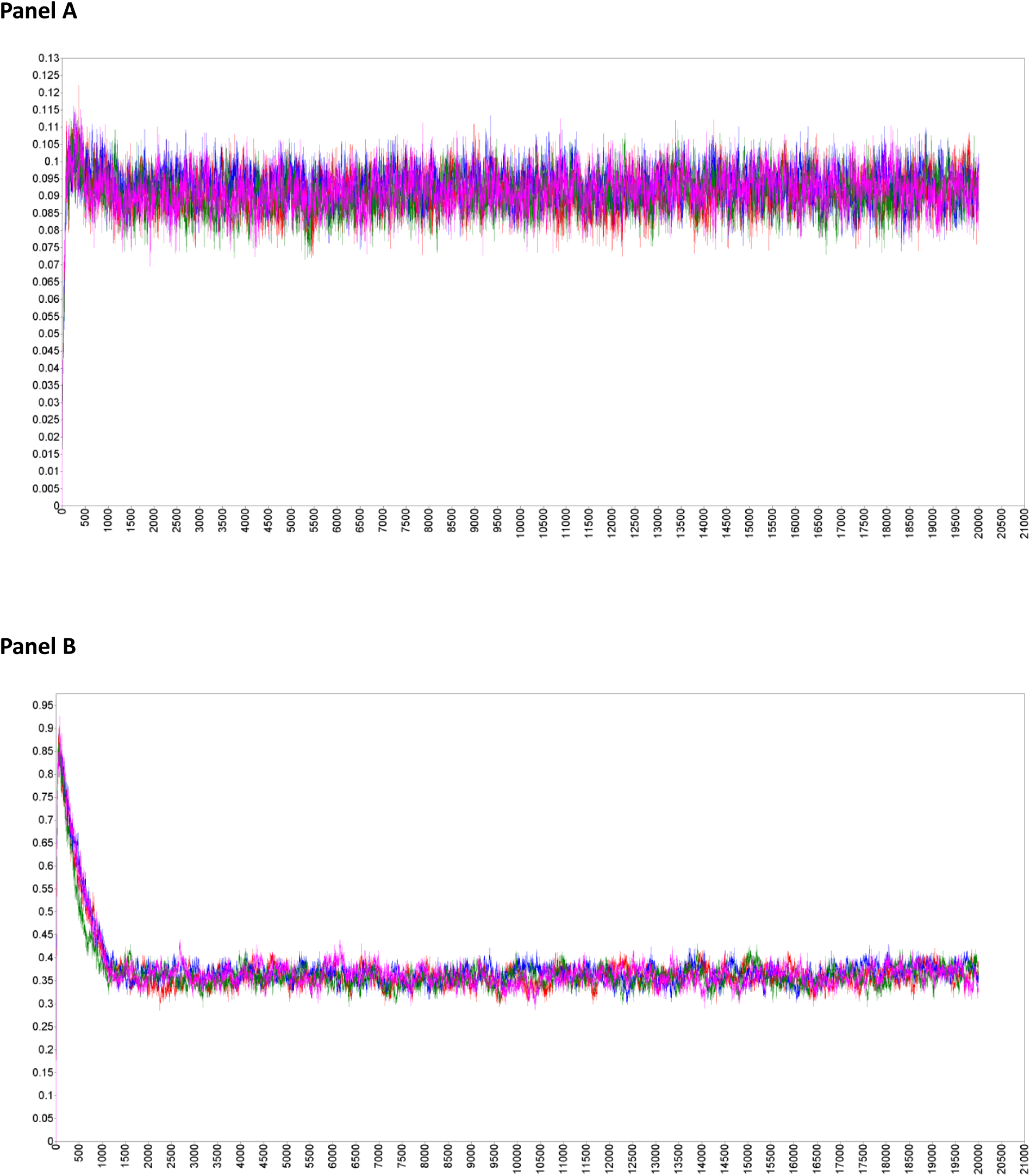
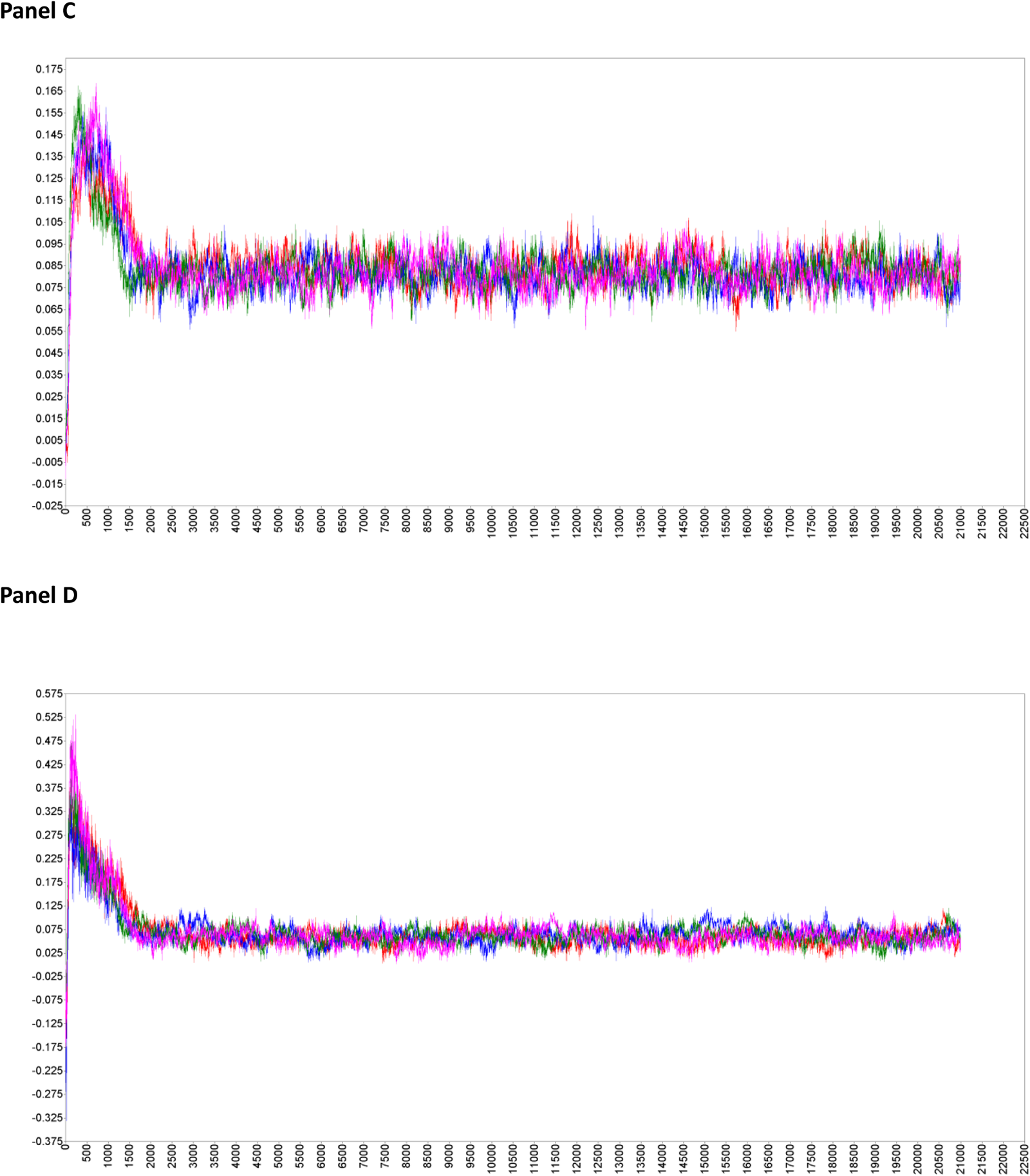

## Appendix E: Results of the sensitivity analysis of auto-regressive and cross-lagged effects in the total sample and analytic samples based on Bayesian mixed regression models

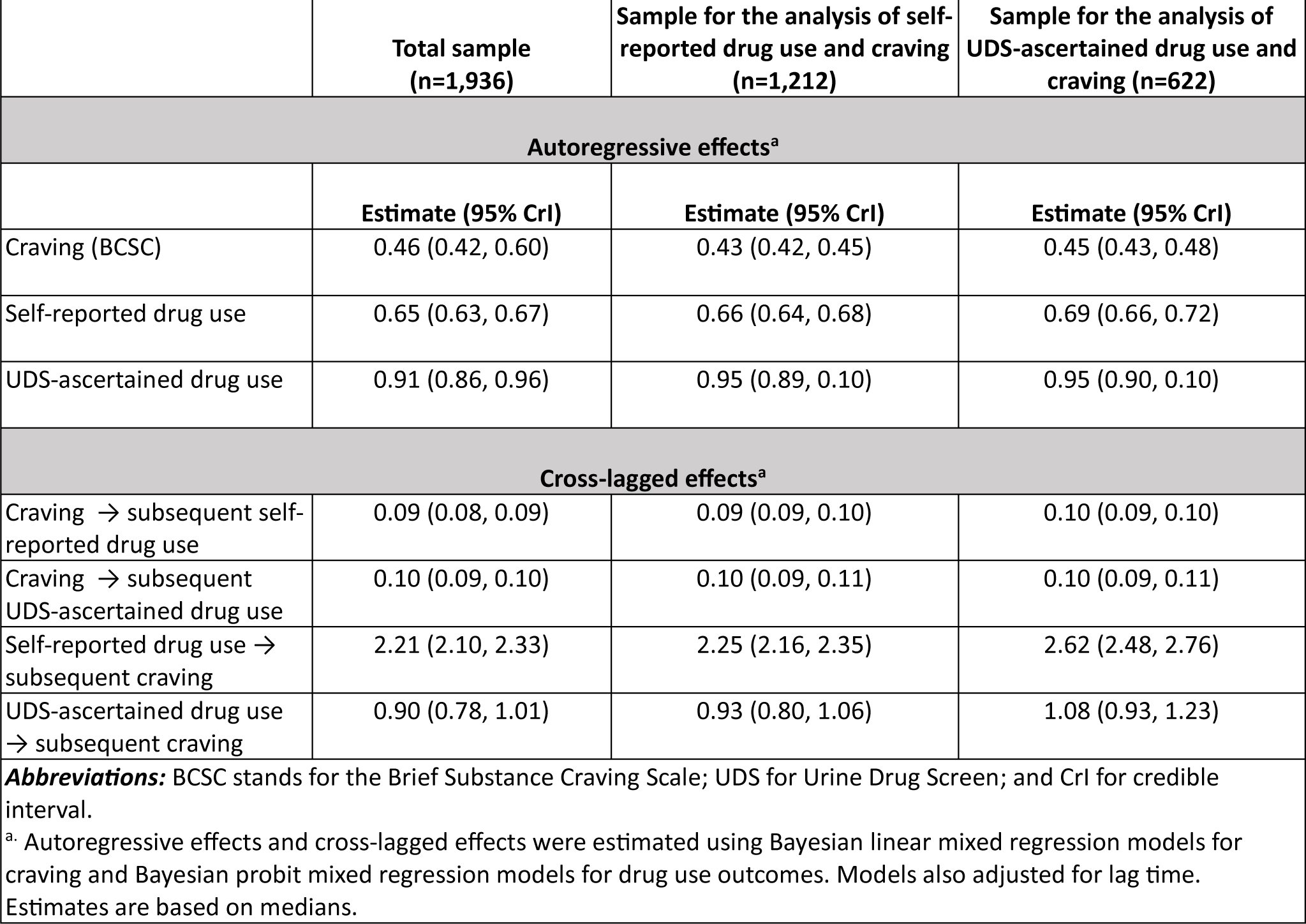

## Appendix F: Results of the Residual Dynamic Structural Equation Modeling (R-DSEM) analyses of the relationship between self-reported drug use and craving as well as urine drug screen (UDS)-ascertained drug use and craving in the course of randomized controlled trials of pharmacotherapies for stimulant use disorders with individual RCT dummy indicators as covariates

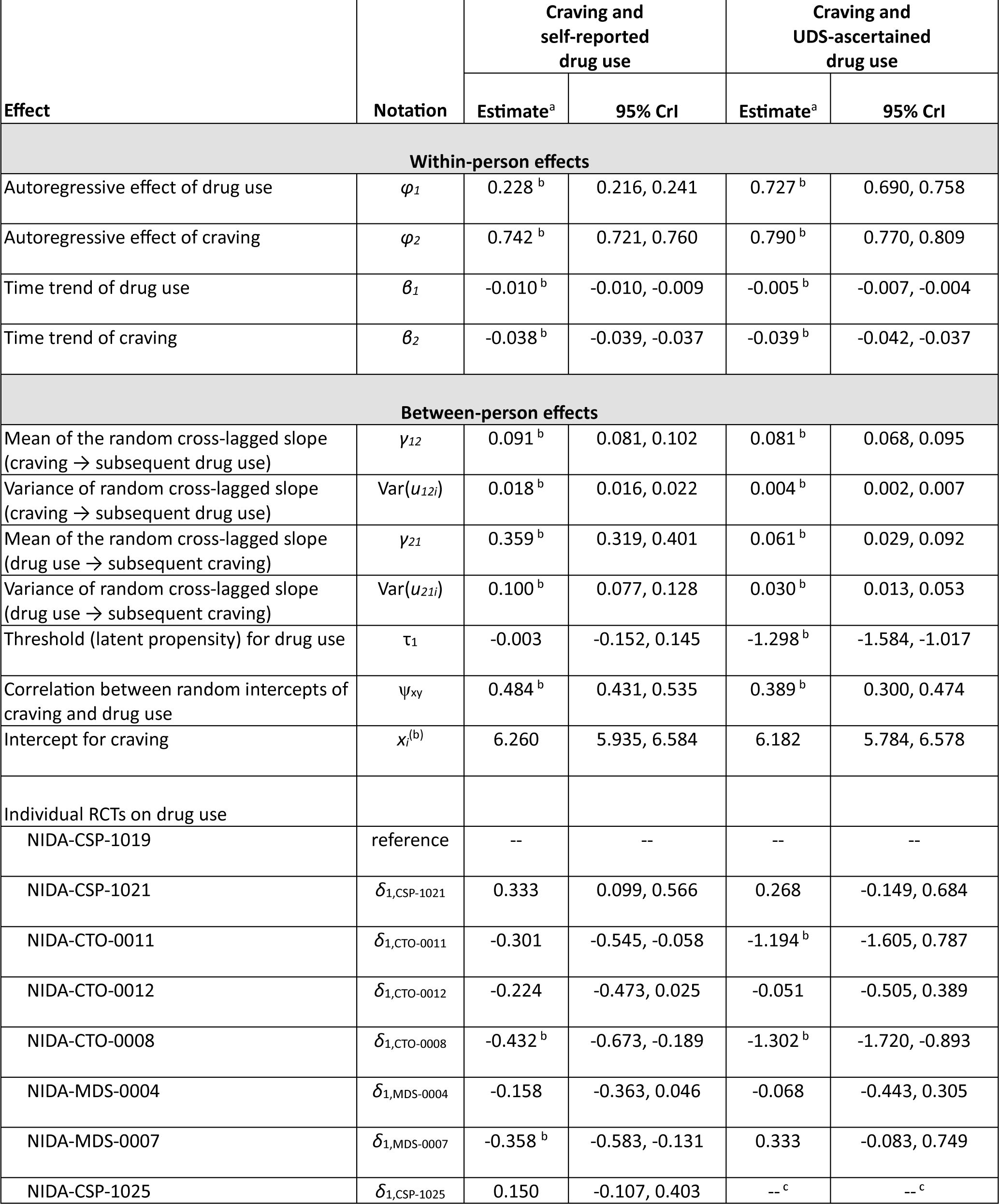
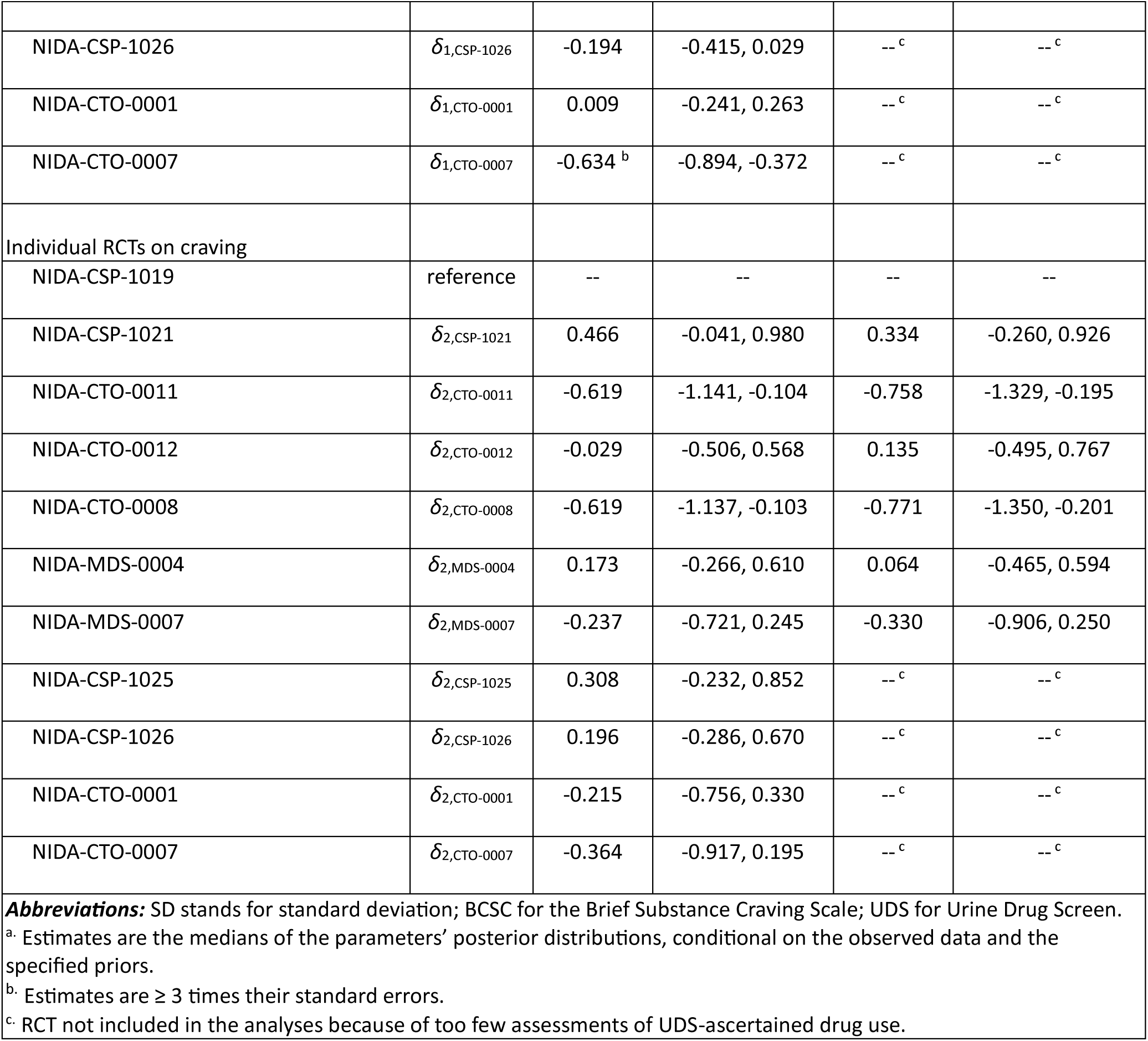

